# US Pediatric Drowning Trends: A System Dynamics Scoping Model Based on Global Burden of Disease (GBD) Estimates

**DOI:** 10.1101/2025.10.12.25337081

**Authors:** Brian J. Biroscak, Grace Kim, Sarah D. Ronis, Tracy E. McCallin, Barbara M. Garza Ornelas, Robinson Salazar Rua

**Affiliations:** Center for Community Health Integration Case Western Reserve University School of Medicine Cleveland, OH, USA; Division of Pediatric Hospital Medicine UH Rainbow Babies and Children’s Hospital Cleveland, OH, USA; Department of Pediatrics University of Florida College of Medicine Gainesville, FL, USA

## Abstract

**Background:** Pediatric drowning remains a leading cause of unintentional injury death globally. Current prevention research primarily focuses on single interventions, often failing to account for the complex interactions and feedback mechanisms that drive population trends. This study demonstrates the utility of system dynamics (SD) modeling as a scoping tool to construct and test causal feedback hypotheses.

**Methods:** A scoping model was developed to simulate the drivers of pediatric drowning epidemiology. Modeling was based on historical drowning morbidity and mortality estimates of one US state from the Global Burden of Disease (GBD) initiative (1990 to 2021), supplemented by secondary literature. Key feedback loops—including the hypothesized intergenerational transfer of swimming competency and the impact of perceived risk on water supervision and prevention investments—were explicitly mapped within the model structure.

**Results:** The model successfully replicated the non-constant decline in pediatric drowning deaths observed in the state-specific GBD data: 50.6 per 100,000 in 1990 versus 25.7 per 100,000 in 2021. Analyses identified the balancing feedback loop (B1) of “Reduced Risk Perception” as a dominant structure. This causal mechanism illustrates that sustained success in mortality reduction can inadvertently lower *Perceived Prevalence*, subsequently reducing *Public Concern* and prevention investments, thereby slowing the overall rate of decline.

**Conclusion:** This study demonstrates that SD modeling is a powerful and accessible tool for testing aggregate, long-term hypotheses regarding unintentional drowning trends. It provides a framework for designing integrated, dynamic prevention strategies that account for the system tendency to adapt and introduce counter-intuitive outcomes.

**WHAT IS ALREADY KNOWN ON THIS TOPIC:** Pediatric drowning rates have decreased due to several hypothesized measures, including improved pool fencing, increased parental supervision, and expanded access to swimming lessons. However, the main scientific knowledge gap is a lack of understanding regarding how these individual interventions interact over time within a complex social system. Specifically, it remains unclear how success in one area (e.g., mortality reduction) influences the effectiveness or sustainability of other measures (e.g., public vigilance and funding).

**WHAT THIS STUDY ADDS:** By integrating Global Burden of Disease (GBD) estimates into a system dynamics framework, this study identifies the causal structures driving observed mortality trends. We identify a “success-bred complacency” dominant causal mechanism that explains the non-linear plateaus and reversals often seen in long-term injury data.

**HOW MIGHT THIS STUDY AFFECT RESEARCH, PRACTICE, OR POLICY:** System dynamics modeling allows for studying shifts in dominant causal feedback mechanisms over time. Future work to disaggregate model trends by pediatric group characteristics (e.g., race and ethnicity) is necessary to inform strategies that further prevent pediatric unintentional drowning.

## Introduction

Unintentional drowning^1^ remains a leading cause of injury-related mortality among children and adolescents.^2^ In the United States, drowning ranks among the top three causes of unintentional injury deaths for individuals aged 29 years and younger.^3^ Although pediatric drowning mortality has declined in recent decades, these trends are non-linear,^4^ and significant disparities persist.^5^ Global Burden of Disease (GBD) initiative data^6^ provide a valuable record of these patterns.^7^

Traditional prevention research reflects linear causal thinking, which may limit understanding of drowning’s multifaceted nature.^8^ The American Academy of Pediatrics identifies five evidence-based prevention strategies: barriers, supervision, life jacket use, water competency, and CPR training.^9^ However, drowning risk exists within a complex adaptive system where single-strategy impacts can be counter-intuitive due to time delays and feedback loops. Successful prevention may lead to diminished risk perception,^10^ subsequently reducing resource allocation. Such hypotheses have not been systematically tested.

System dynamics (SD) modeling serves as a tool for understanding causal structures in complex phenomena,^11^ including drowning prevention. This paper presents a scoping model^12^ demonstrating its utility in synthesizing prevention knowledge and leveraging GBD mortality estimates (ages 0-19 years). The objective is to provide a framework for creating expansive research models informing integrated prevention strategies.

## Methods

### Model Conceptualization

The scoping model synthesizes and tests causal feedback hypotheses regarding pediatric drowning trends. The SD model includes five primary stocks: *Individuals at Risk*, *Skilled Swimmers*, *Drowning Incidents*, *Investments in Prevention*, and *Perceived Prevalence*. Dynamic flows (e.g., *Incidence from Domestic/Residential Drowning*) govern the system’s rate of change (**Table 1**).

**Table 1.** System Dynamics Scoping Model Main Variables and Descriptions.

| Variable Type | Variable Name | Description | Unit |
| --- | --- | --- | --- |
| Stock | Individuals at Risk | The population (ages 0–19) exposed to unintentional drowning risk (e.g., non-swimmers or unsupervised). | People |
| Stock | Skilled Swimmers | The population (ages 0–19) who possess water competency, acting as a protective factor. | People |
| Stock | Drowning Incidents | The accumulated population of non-fatal drowning events requiring intervention/recovery. | People |
| Stock | Investments in Prevention | The resources allocated over time to prevention efforts (e.g., funding for lessons, barriers). | USD/Year |
| Stock | Perceived Prevalence | The subjective perception of drowning prevalence within society, which adjusts gradually to actual prevalence with a time delay. | Dimensionless |
| Flow | Skill Gains | The rate at which *Individuals at Risk* transition to *Skilled Swimmers* (e.g., through lessons). | People/Year |
| Flow | Skill Losses | The rate at which water competency is lost due to factors like atrophy or lack of practice. | People/Year |
| Flow | Drowning Deaths | The rate at which *Drowning Incidents* result in fatal outcomes. | People/Year |

The model distinguishes *Natural/Open Water* (lakes, rivers, oceans) from *Domestic/Residential* (pools, bathtubs) drowning. Residential drowning shows higher intervention elasticity (through barriers and supervision), whereas natural water drowning depends more on water competency. This distinction captures differential prevention strategy effectiveness.

The structure formalizes two dynamic hypotheses. First, a reinforcing feedback loop (R) where successful prevention leads to higher water competency, driving mortality down. Second, a balancing loop (B) where prevention success inadvertently lowers perceived risk, reducing motivation for future investment and slowing mortality decline.

### Data and Implementation

GBD estimation includes compartmental modeling.^13^ The primary reference data were unintentional drowning mortality rates among persons aged 0–19 in Ohio (deaths per 100,000), derived from GBD estimates (1990–2021). The model was built using Stella Architect.^14^ Testing focused on behavior reproduction—replicating the non-constant historical decline. Internal structure was confirmed by assessing logical consistency against secondary literature and professional knowledge.

## Results

### Behavior Reproduction and Dominant Loop Identification

The calibrated model successfully replicated Ohio’s pediatric drowning mortality trend from 1990 to 2021, decreasing from 50.6 to 25.7 deaths per 100,000 (**Figure 1**). This was accompanied by simulated increases in *Investments in Prevention Resources* and *Skilled Swimmers*. Dynamic analysis identified one dominant balancing loop (**Figure 2**) explaining 40-50% of system behavior (**Figure 3**). This loop (B1), “Reduced Risk Perception,” primarily drives the long-term decline.

### Mechanism of the Balancing Loop (B1)

B1 describes continuous, adaptive societal response:

1. **Success and Delayed Perception:** As actual *Drowning Prevalence* declines through prevention, *Perceived Prevalence* adjusts downward with delay, reflecting slow updating of public awareness.
2. **Complacency Effect:** Declining *Perceived Prevalence* reduces *Public Concern*, modulating growth of *Investments in Prevention Resources*. Because perception lags reality, residual concern sustains—but does not accelerate—continued investment.
3. **Investment and Skill Gain:** Sustained (but growth-limited) *Investments* increase the *Skill Gain Rate* (through lessons and education), expanding *Skilled Swimmers*.
4. **Mortality Reduction:** Growing *Skilled Swimmers* reduces *Individuals at Risk*, declining incidence from both *Natural/Open Water* and *Domestic/Residential* activities, further reducing *Drowning Prevalence*.

This balancing loop creates goal-seeking behavior: as success reduces prevalence, complacency resists rapid change. The system settles into sustained but decelerating decline—explaining the non-linear GBD trajectory. Loop B1’s dominance (40-50%, Figures 2-3) demonstrates that complacency, rather than resource constraints, primarily limits mortality reduction pace.

## Discussion

This SD scoping model provides a framework for understanding non-linear dynamics in pediatric drowning mortality decline. Recently, some jurisdictions report reversals following COVID-19, possibly from reduced swim lesson access and altered supervision.^15^ The BI loop dominance offers insight transcending single-intervention evaluation.

The model suggests sustained reduction is not simply summed individual efforts, but dynamic, self-correcting societal processes. B1 formalizes that ‘success breeds both progress and challenges’. As drowning declines, reduced prevalence lowers *Perceived Prevalence*, reducing *Public Concern* and dampening prevention investment pressure. This complacency effect creates resistance to rapid decline, explaining non-constant deceleration.

This finding carries implications for policy and practice. First, it underscores need for sustained funding. Investment is driven by time-delayed feedback, not policy impulse. Programs must persist for decades to maintain system pressure.

Second, the model supports a behavioral hypothesis explored in other risk domains: inverse relationship between mitigation success and risk perception. This aligns with the Social Amplification of Risk Framework—successful risk dampening can lead to complacency.^16^ To counteract inherent system tendency toward complacency, communication strategies must be dynamic, continuously adjusting based on *Perceived Prevalence* decline. This involves shifting from “high-urgency” during higher-risk periods to “maintenance” messaging addressing subtle risks (skill atrophy, supervision lapses) and emphasizing persistent vigilance, intentionally stabilizing B1.

### Limitations

As a scoping model, this study is limited. Parameter estimates relied on literature synthesis and calibration to aggregate Ohio GBD data, restricting generalizability. The model operates at an aggregate level, not accounting for race, socioeconomic status, or geographical variables driving disparities. Future research must expand this framework, testing against localized data for targeted, equity-focused interventions.

Calibration to a single geography (Ohio) is notable. However, preliminary analysis of GBD modes for Great Lake states suggests similar system archetypes, indicating B1 may function as generic structure in pediatric injury—a fundamental societal feedback operating across jurisdictions. Future work should involve multi-state validation determining if ‘success-bred complacency’ is regional or national, and whether it varies across risk environments (*Natural/Open Water* versus *Domestic/Residential*).

## Conclusion

This scoping model demonstrated SD utility for analyzing long-term, non-linear pediatric drowning mortality decline. By mapping system structure, analysis revealed “Reduced Risk Perception” balancing loop as dominant driver, explaining why mortality declines at sustained yet non-constant pace. This argues against linear projection evaluations.

Policymakers should recognize drowning prevention as an adaptive, complex challenge driven by time-delayed feedback. We seek to engage the injury research field to co-develop this scoping model, converting it into validated regional research models.

Effective long-term injury prevention requires designing integrated interventions accounting for counter-intuitive outcomes and establishing mechanisms for sustained resource investment preventing risk complacency.

## Data Availability

All data used in this study are publicly available. Historical drowning mortality estimates (1990-2021) were obtained from the Global Burden of Disease Study 2021 (GBD 2021), Institute for Health Metrics and Evaluation (IHME), available at
https://vizhub.healthdata.org/gbd-results/. The system dynamics model structure, equations, and calibrated parameters are available from the corresponding author upon reasonable request. The Stella Architect model file (.stmx format) is
available upon request for researchers with access to compatible software.

## Data availability statement

Model calibration utilized publicly available estimates from the Global Burden of Disease Study 2021 (GBD 2021); specific model equations are available upon request from the corresponding author.

## Ethics statements

### Patient consent for publication

Not applicable.

### Ethics approval

Not applicable.

## Acknowledgements

The authors thank the Global Burden of Disease (GBD) initiative and the Institute for Health Metrics and Evaluation (IHME) for providing the foundational estimates used in this study. In accordance with GBD publication policy, a request for review and circulation has been submitted to the GBD Secretariat. This study was conducted as a secondary analysis by GBD Collaborators. We invite readers interested in applying the SD methodology to specific, localized drowning data to contact the corresponding author to facilitate the conversion of this scoping model into regional research models.

## Footnotes

### Funding

This study did not receive any funding.

### Competing interests

The authors have declared no competing interests.

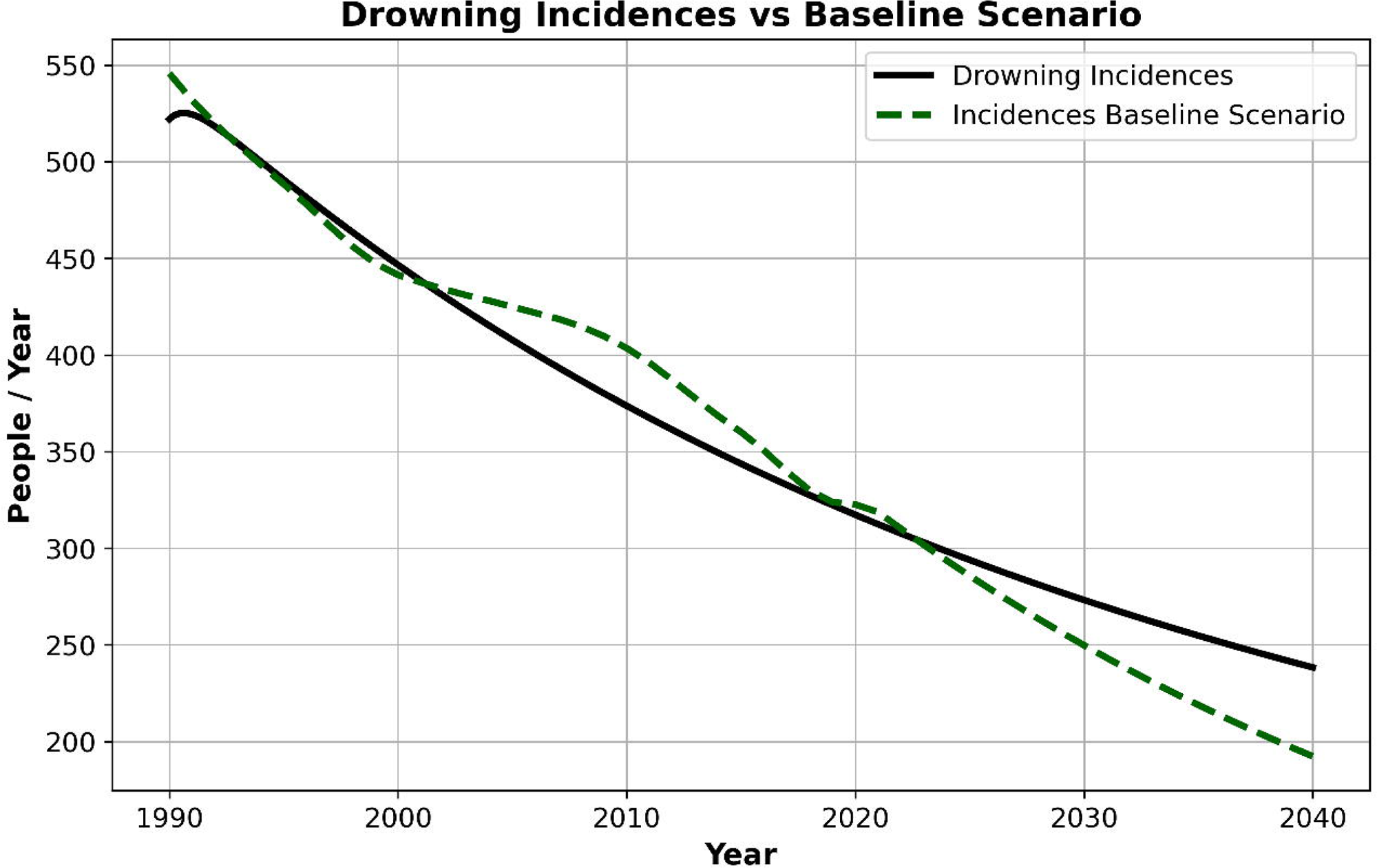

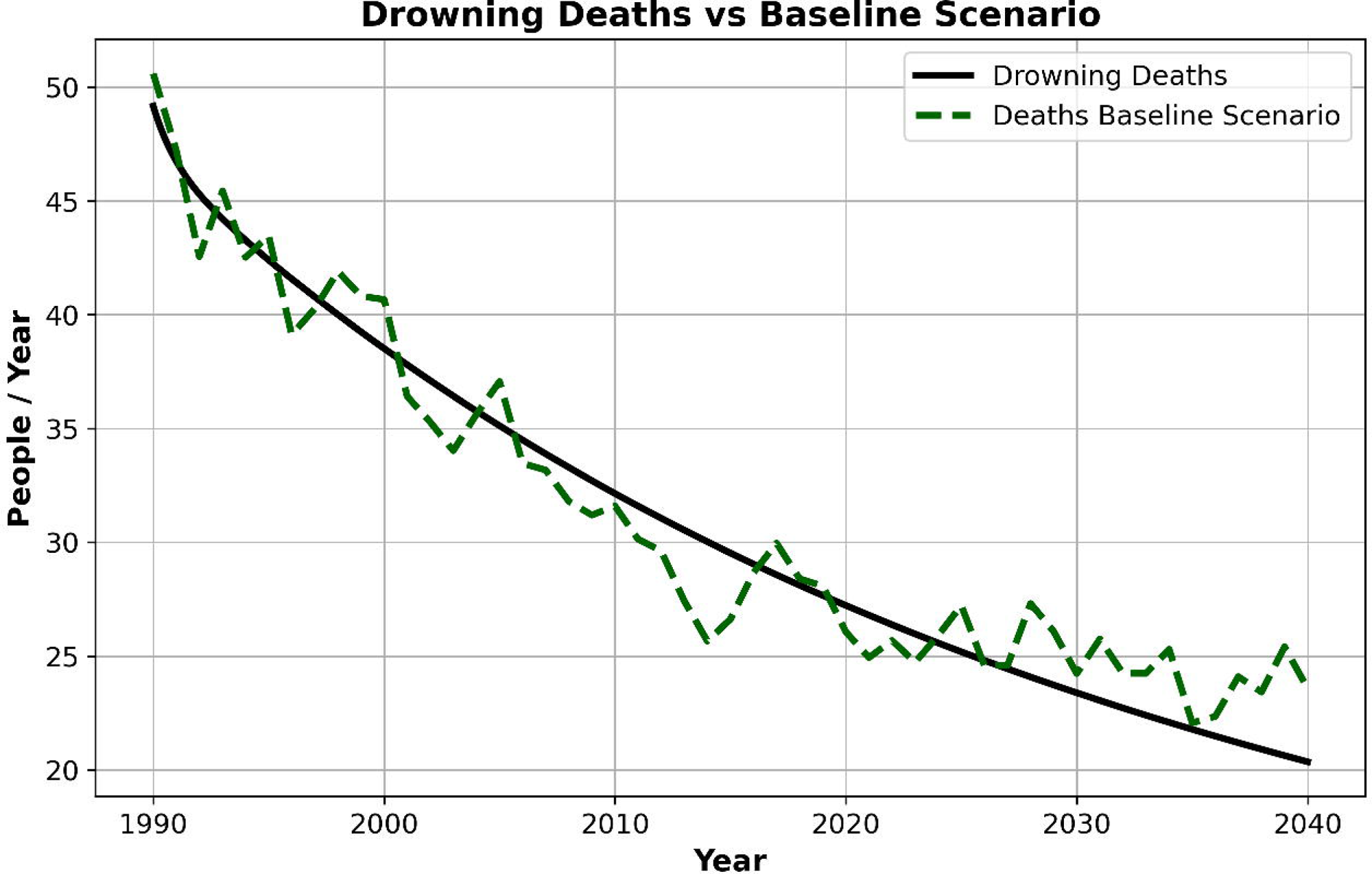

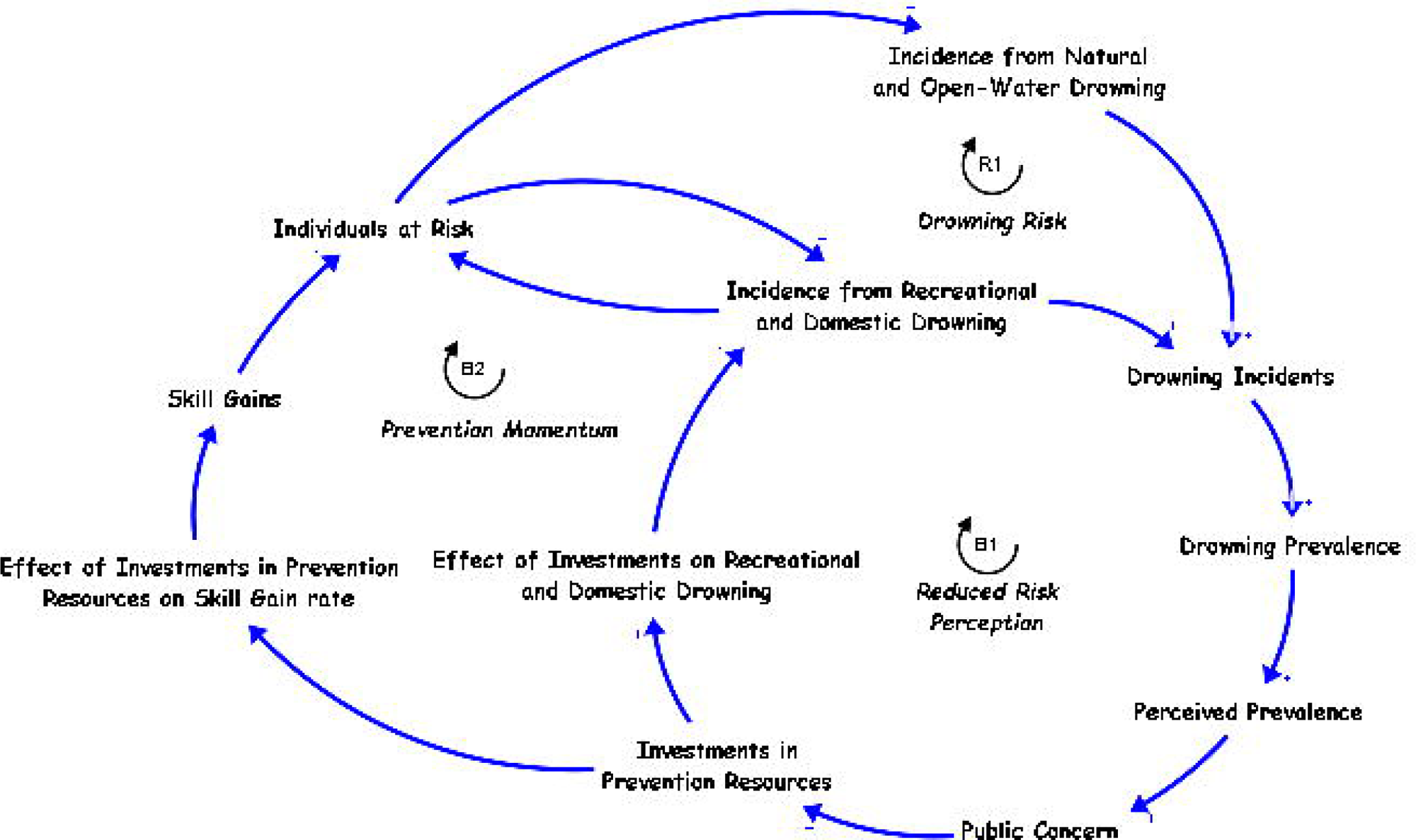

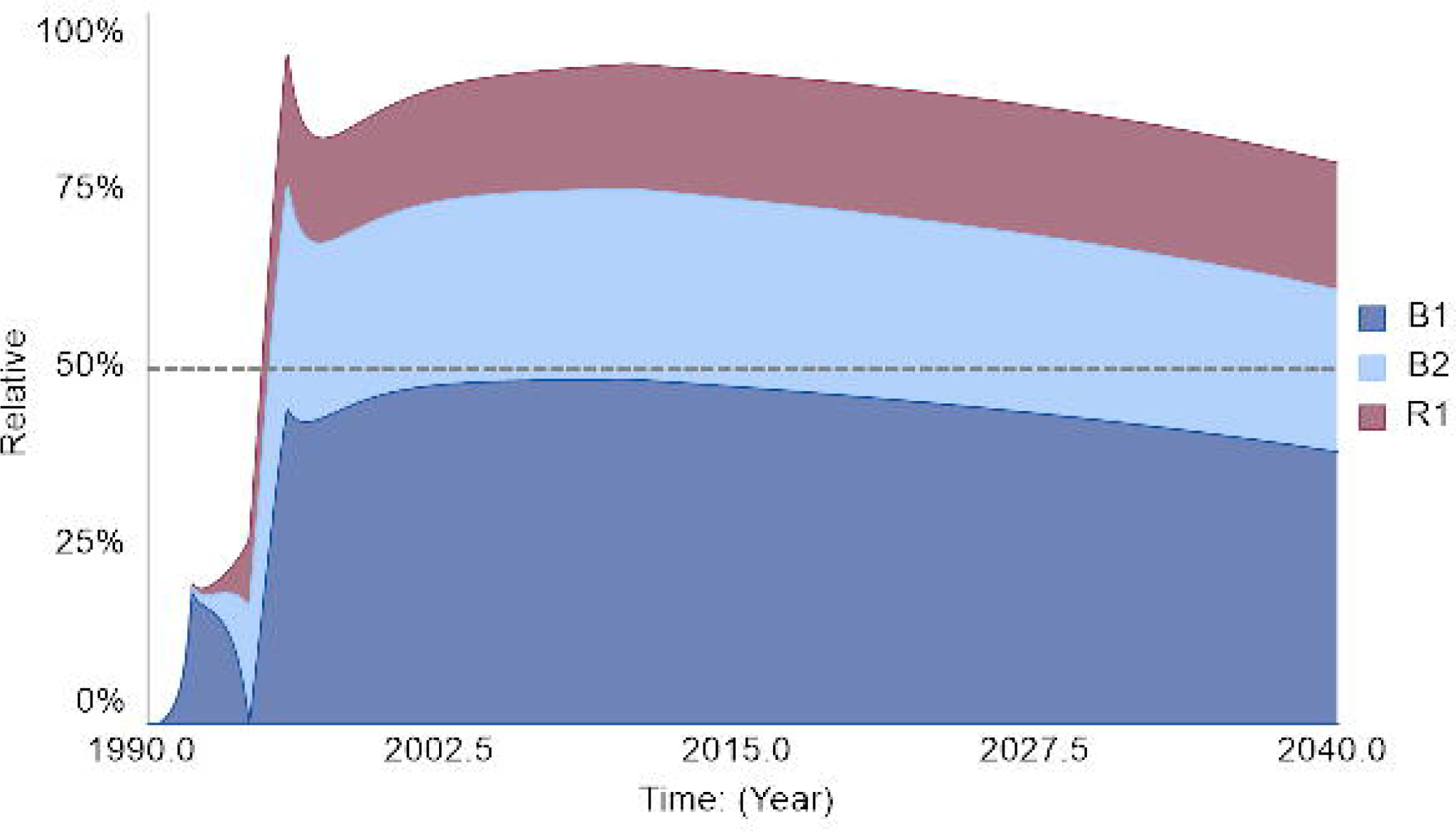

## Notes

### Author Declarations

The study used ONLY openly available data that were originally located at: Global Burden of Disease Results Tool, Institute for Health Metrics and Evaluation (IHME), University of Washington. https://vizhub.healthdata.org/gbd-results/ (accessed January 2025). No registration, application, or institutional approval was required to access these publicly available aggregate mortality estimates.

### Summary of Updates

**Update [12/31/2025]:** Major revisions from the original preprint include: corrected model mechanism description for B1 loop; added quantitative loop dominance metrics; streamlined text to meet word limit for BMJ publication, Injury Prevention.

