## Supplemental File 1 for "US Pediatric Drowning Trends: A System Dynamics Scoping Model Based on Global Burden of Disease (GBD) Estimates"

STRESS-SD Checklist

### 1. OBJECTIVES

#### 1.1 Purpose of the model

**Location in manuscript:** Introduction, paragraph 3

**Response:** This scoping model demonstrates the utility of system dynamics methodology for testing aggregate, long-term causal hypotheses regarding pediatric unintentional drowning trends. The model synthesizes existing prevention knowledge and leverages Global Burden of Disease (GBD) estimates to provide a framework for creating more expansive research models that can inform integrated and dynamic drowning prevention strategies.

#### 1.2 Model Outputs

**Location in manuscript:** Results; Table 1; Figures 1-3

**Response:** The primary outcome variables reported are:

1. **Drowning Deaths** (People/Year) - Fatal outcomes calculated as: Drowning Incidents × Death Rate (0.0494)
2. **Drowning Incidence** (People/Year) - Sum of two flow variables:
   - Incidence from Natural/Open Water = Individuals at Risk × Risk Rate from Natural/Open Water Activities / IP3
   - Incidence from Domestic/Residential = Individuals at Risk × Risk Rate from Recreational/Domestic / IP3
3. **Drowning Prevalence** (Dimensionless) - Calculated as: Drowning Incidents / Total Population (100,000)
4. **Skilled Swimmers** (People) - Stock variable tracking water-competent population
5. **Investments in Prevention Resources** (USD/Year) - Stock variable tracking resource allocation

All outputs are reported as time series from 1990-2021 (historical) and projections to 2040.

#### 1.3 Experimentation Aims

**Location in manuscript:** N/A - This is a scoping model, not policy analysis

**Response:** This scoping model was developed for behavior reproduction and hypothesis testing, not policy experimentation. No user-defined policy experiments or optimization were conducted. The model includes infrastructure for future sensitivity analysis through intervention parameters (IP1-IP6 with corresponding effect sizes ES1-ES6), but these were not systematically varied in this study. Future research will use this framework for policy analysis across different jurisdictions and demographic groups.

### 2. LOGIC

#### 2.1 Base model overview diagram

**Location in manuscript:** Figure 2 (simplified causal loop diagram)

**Response:** Provided. The model includes:

- Stock-flow diagram showing all five stocks, flows, and auxiliary variables (available upon request)
- Simplified causal loop diagram highlighting three key feedback loops: B1 (Reduced Risk Perception), B2 (Prevention Momentum), and R1 (Drowning Risk) (Figure 2)

#### 2.2 Base model logic

**Location in manuscript:** Methods (Model Conceptualization); Results (Mechanism of B1)

**Response:** The model is structured around three primary feedback loops:

#### B1 - Reduced Risk Perception (Balancing, Dominant 40-50%):

- Drowning Prevalence ↓ → Perceived Prevalence ↓ (delayed) → Public Concern ↓ → Investments ↓ → Skill Gains slower → Prevalence decline slows
- This complacency mechanism creates goal-seeking behavior, explaining sustained but decelerating mortality decline

#### B2 - Prevention Momentum (Balancing):

- Individuals at Risk → Skill Gains → Skilled Swimmers → Skill Losses (atrophy) → back to Individuals at Risk
- Represents intergenerational transfer and loss of water competency

#### R1 - Drowning Risk (Reinforcing):

- Individuals at Risk → Drowning Incidence → Drowning Incidents (stock) → through various pathways back to risk exposure
- Represents the self-amplifying nature of drowning risk without intervention

#### 2.3 Scenario logic

**Location in manuscript:** N/A

**Response:** Not applicable. This scoping model presents a single calibrated base case. No alternative scenarios, policies, or interventions were tested. The model structure includes intervention points (IP1-IP6) for future scenario analysis, but these remain at baseline (multiplier = 1.0) in this study.

#### 2.4 Algorithms

**Location in manuscript:** Methods (Data and Implementation)

**Response:** No complex scheduling or manual process algorithms were required. Standard system dynamics integration methods were used. The model employs:

- First-order exponential delays (represented through stock-flow structures)
- Power law formulation for societal pressure response: Indicated Investments = Initial Investments × (Public Concern / INIT(Public Concern))^0.812
- Linear proportional relationships for investment effects on skill gains and risk rates

#### 2.5 Components

2.5.1 **Stocks/Levels**

**Location in manuscript:** Table 1

#### Response:

| **Stock** | **Initial Value** | **Units** | **Inflows** | **Outflows** | **Description** |
| --- | --- | --- | --- | --- | --- |
| Individuals at Risk | 19,066 | People | Drowning Recoveries; Skill Losses | Skill Gains; Incidence (Natural/Open); Incidence (Domestic/Residential) | Population (ages 0-19) exposed to drowning risk |
| Skilled Swimmers | 1,000 | People | Skill Gains | Skill Losses | Population (ages 0-19) with water competency |
| Drowning Incidents | 1,000 | People | Incidences (Natural/Open); Incidences (Domestic/Residential) | Drowning Deaths; Drowning Recoveries | Accumulated non-fatal drowning events |
| Investments in Prevention | 1,000,000 | USD/Year | Change in Investments | (none - represents flow rate itself) | Resources allocated to prevention efforts |
| Perceived Prevalence | INIT(Prevalence) | Dimensionless | Change in Perceived Prevalence | (none) | Society's subjective perception of drowning prevalence |

2.5.2 **Flows/Rates**

**Location in manuscript:** Table 1

#### Response:

| **Flow** | **Units** | **Equation** | **Role** |
| --- | --- | --- | --- |
| Skill Gains | People/Year | Individuals at Risk × Skill Gain Rate × IP1 | Transition from at-risk to skilled |
| Skill Losses | People/Year | Skilled Swimmers × 0.1147 / IP2 | Water competency atrophy |
| Incidence from Domestic/Residential | People/Year | Individuals at Risk × Risk Rate (Domestic) / IP3 | Drowning events in pools/homes |
| Incidence from Natural/Open Water | People/Year | Individuals at Risk × Risk Rate (Natural) / IP3 | Drowning events in lakes/rivers |
| Drowning Deaths | People/Year | Drowning Incidents × 0.0494 | Fatal outcomes |
| Drowning Recoveries | People/Year | Drowning Incidents × 0.543 × IP4 | Non-fatal incident resolution |
| Change in Investments | USD/Year/Year | (Indicated Investments - Investments) × 0.2265 × IP6 | Adjustment of prevention funding |
| Change in Perceived Prevalence | 1/Year | (Prevalence - Perceived Prevalence) × 0.995 × IP5 | Delayed perception adjustment (information delay ~1 year) |

2.5.3 **Constants / Converters / Auxiliary variables Location in manuscript:** Methods section

#### Response:

| **Variable** | **Value** | **Units** | **Equation/Description** |
| --- | --- | --- | --- |
| Total Population | 100,000 | People | Reference population for rate calculations |
| Risk Percentage | 0.1906 | Dimensionless | Proportion of population at risk (calibrated) |
| Initial Investments | 1,000,000 | USD/Year | Baseline prevention investment level |
| Base Skill Gain Rate | 0.1954 | 1/Year | Calibrated rate of skill acquisition |
| Base Risk Rate (Residential) | 0.4646 | 1/Year | Calibrated base rate for residential drowning |
| Base Risk Rate (Open Water) | 0.0697 | 1/Year | Calibrated base rate for open water drowning |
| Death Rate | 0.0494 | Dimensionless | Case fatality rate (calibrated) |
| Recovery Rate | 0.5430 | Dimensionless | Non-fatal incident resolution rate |
| Skill Loss Rate | 0.1147 | 1/Year | Water competency atrophy rate |
| Investment Adjustment Rate | 0.2265 | 1/Year | Speed of investment response (time constant ~4.4 years) |
| Perception Adjustment Rate | 0.995 | 1/Year | Speed of perception update (time constant ~1.0 year) |
| Public Concern Elasticity | 0.812 | Dimensionless | Power law exponent for pressure-investment relationship |
| Skill Gain Rate | Base Rate × Effect of Investments | 1/Year | Investments increase skill acquisition |
| Risk Rate (Residential) | Base Rate / Effect of Investments | 1/Year | Investments reduce residential drowning risk |
| Risk Rate (Open Water) | Base Rate / Effect of Investments | 1/Year | Investments reduce open water drowning risk |
| Effect of Investments on Skills | Investments / Initial Investments | Dimensionless | Proportional multiplier (>1 increases skills) |

**Variable	Value	Units	Equation/Description**

| Effect of Investments on Risk | Initial Investments / Investments | Dimensionless | Inverse multiplier (>1 investment reduces risk) |
| --- | --- | --- | --- |
| Public Concern | Perceived Prevalence | Dimensionless | Direct proportionality (higher perception = higher concern) |
| Indicated Investments | Initial × (Concern/INIT(Concern))^0.812 | USD/Year | Goal-seeking investment level |
| Prevalence | Drowning Incidents / Total Population | Dimensionless | Actual drowning rate |
| IP1 through IP6 | 1.0 (baseline) | Dimensionless | Intervention parameters (reserved for future scenarios) |
| ES1 through ES6 | 0.0 (baseline) | Dimensionless | Effect sizes (reserved for future scenarios) |

#### 2.5.4 Graphical functions/lookup tables Location in manuscript: N/A

**Response:** The model includes three graphical functions for baseline comparison data (not used in model calculations, only for visualization):

- Incidence Baseline Scenario (TIME) - Historical GBD incidence data
- Deaths Baseline Scenario (TIME) - Historical GBD mortality data
- Prevalence Baseline Scenario (TIME) - Calculated historical prevalence

These are lookup tables populated with empirical GBD data for Ohio (1990-2021) used solely for graphical comparison.

#### 2.5.5 Sources and Sinks Location in manuscript: simulation model available upon request

**Response:**

- **Infinite Source:** Cloud symbol at origin of Skill Gains flow (representing new cohorts entering the at-risk population through birth/aging into 0-19 age range)
- **Infinite Sink:** Cloud symbol at terminus of Drowning Deaths flow (representing mortality exit from the modeled population)
- **Model Boundary:** The model bounds the pediatric population (ages 0-19) and does not explicitly model aging out of this cohort, treating it as a closed population for the 31-year historical period

### 3. DATA

#### 3.1 Data sources

**Location in manuscript:** Methods (Data and Implementation); References 6-7

**Response:**

**Primary Data Source:**

- **Global Burden of Disease Study 2021 (GBD 2021)**
  - Publisher: Institute for Health Metrics and Evaluation (IHME)
  - Data type: Publicly available aggregate mortality estimates
  - Geographic scope: Ohio, USA
  - Population: Ages 0-19 years
  - Variables: Unintentional drowning deaths per 100,000 population
  - Date range: 1990-2021 (annual)
  - Sample size: 32 annual data points
  - Use: Model calibration and validation (behavior reproduction)
  - Reference: Global Burden of Disease Collaborative Network. Global Burden of Disease Study 2021 (GBD 2021). Seattle, WA, USA: IHME; 2025.

#### Secondary Literature Sources:

- Parameter estimation relied on synthesis of published literature on:
  - Swimming lesson effectiveness
  - Water competency retention and skill atrophy
  - Drowning case fatality rates
  - Recovery rates from non-fatal drowning incidents
  - Prevention investment trends

#### Stakeholder Input:

- Clinical expertise from pediatric hospital medicine physicians (co-authors) informed plausible ranges for parameters and model structure validation

#### 3.2 Pre-processing

**Location in manuscript:** Methods (Data and Implementation)

#### Response:

**GBD Data:**

- No interpolation required (annual data complete for 1990-2021)
- No outliers removed (all data points used)
- Data converted from deaths per 100,000 to absolute numbers using reference population of 100,000 for modeling purposes

#### Literature-derived Parameters:

- Where parameter ranges were available from literature, midpoint values were selected as starting points for calibration
- No distributional fitting was performed (deterministic model)

#### 3.3 Input parameters

**Location in manuscript:** Methods; Section 2.5.3 above; Data Availability Statement

**Response:**

**Base Case Parameters (All Calibrated to Ohio GBD Data 1990-2021):**

| **Parameter** | **Value** | **Units** | **Source** | **Calibration Method** |
| --- | --- | --- | --- | --- |
| Initial Individuals at Risk | 19,066 | People | Derived: Total Population × Risk Percentage | Optimization |
| Risk Percentage | 0.1906 | Dimensionless | Literature + Calibration | Optimization to fit initial conditions |
| Initial Skilled Swimmers | 1,000 | People | Assumed (low initial competency) | Assumed |
| Initial Drowning Incidents | 1,000 | People | Assumed | Assumed |
| Initial Investments | 1,000,000 | USD/Year | Assumed (indexed value) | Assumed |
| Base Skill Gain Rate | 0.1954 | 1/Year | Literature + Calibration | Least-squares optimization |
| Skill Loss Rate | 0.1147 | 1/Year | Literature (competency atrophy studies) + Calibration | Least-squares optimization |
| Base Risk Rate (Domestic) | 0.4646 | 1/Year | Calibration | Least-squares optimization |
| Base Risk Rate (Natural) | 0.0697 | 1/Year | Calibration | Least-squares optimization |
| Death Rate | 0.0494 | Dimensionless | Calibration to match GBD deaths | Least-squares optimization |
| Recovery Rate | 0.5430 | Dimensionless | Literature + Calibration | Least-squares optimization |
| Investment Adjustment Rate | 0.2265 | 1/Year | Calibration (policy delay ~4-5 years) | Least-squares optimization |
| Perception Adjustment Rate | 0.995 | 1/Year | Calibration (information delay ~1 year) | Least-squares optimization |
| Public Concern Elasticity | 0.812 | Dimensionless | Calibration | Least-squares optimization |

**Time-Dependent Parameters:**

- All intervention parameters (IP1-IP6) and effect sizes (ES1-ES6) are set to baseline values (1.0 and 0.0 respectively) throughout the base case simulation
- No scenarios with time-varying policies were tested in this study

#### Optimization Details:

- Algorithm: Powell's method (available in Stella Architect)
- Objective: Minimize sum of squared errors between model output and GBD data for both drowning incidence and deaths (dual payoff)
- Parameters optimized: Risk Percentage, Total Population (within constraints), and all rate constants listed above
- Constraints: Non-negative parameters; physically plausible ranges based on literature

**No stochastic inputs used** - This is a deterministic model.

#### 3.4 Assumptions

**Location in manuscript:** Methods; Limitations section

#### Response:

**Structural Assumptions:**

1. **Closed Population:** The model treats ages 0-19 as a closed system, not explicitly modeling births, aging out, or migration (assumes demographic stability over 1990-2021 period)
2. **Aggregate Representation:** No disaggregation by race, ethnicity, socioeconomic status, or specific geography within Ohio (acknowledged as key limitation)
3. **Two-Pathway Structure:** Drowning risk separated into only two contexts (Natural/Open Water vs Domestic/Residential), ignoring other potential contexts
4. **Linear Investment Effects:** Effect of investments on skill gains and risk reduction assumed proportional (Effect = Investment Ratio^1.0), though public concern uses non-linear power law
5. **Instantaneous Skill Transfer:** Skill Gains flow represents immediate transition from at-risk to skilled (no learning delay modeled explicitly)
6. **Homogeneous Perception:** Perceived Prevalence represents aggregate societal awareness, not heterogeneous sub-population perceptions
7. **Historical Investment Proxy:** Investments in Prevention stock uses indexed values (starting at

$1M/year) as proxy for aggregate prevention resources, not actual budget data

#### Parameter Value Assumptions (where empirical data unavailable):

1. Initial stock values (Skilled Swimmers = 1,000; Drowning Incidents = 1,000) assumed low to allow model to reach equilibrium during calibration
2. Initial Investments = $1,000,000/year assumed as indexed baseline (relative, not absolute spending)
3. Time delays (investment adjustment = 4-5 years; perception adjustment = 1 year) based on plausible policy and information diffusion timescales, refined through calibration

#### Behavioral Assumptions:

1. Public Concern is directly proportional to Perceived Prevalence (not a more complex utility or risk perception function)
2. Indicated Investments follow power law response to societal pressure with elasticity 0.812 (sub-linear but responsive)
3. Skill atrophy occurs at constant fractional rate (0.1147/year ≈ 8.7-year half-life of competency)

### 4. EXPERIMENTATION

#### 4.1 Initialisation

**Location in manuscript:** Section 2.5.1 above; Model documentation

#### Response:

**Initial Values (Year 1990):**

- Individuals at Risk = 19,066 People
- Skilled Swimmers = 1,000 People
- Drowning Incidents = 1,000 People
- Investments in Prevention = 1,000,000 USD/Year
- Perceived Prevalence = INIT(Drowning Incidents / Total Population) = 0.01 Dimensionless

**No stochastic initialization** - All initial values are deterministic constants.

**Optimization Note:** Risk Percentage and Total Population were allowed to vary during calibration optimization within constraints (Risk % 0.01-1.0; Total Pop 10M-12M) to achieve best fit to GBD data.

#### 4.2 Run length

**Location in manuscript:** Methods

#### Response:

- **Simulation period:** 1990-2040 (50 years)
- **Historical calibration period:** 1990-2021 (31 years)
- **Projection period:** 2022-2040 (19 years)
- **Time units:** Years
- **Time step (DT):** 0.0625 years (Euler integration)

#### 4.3 Estimation approach

**Location in manuscript:** Methods (Data and Implementation); Section 3.3 above

#### Response:

**Model Type:** Deterministic (no stochastic inputs)

- No multiple replications required
- Single model run produces deterministic output trajectory

#### Calibration Method:

- **Algorithm:** Powell's method (gradient-free optimization)
- **Objective Function:** Minimize weighted sum of squared errors
  - Payoff 1: Least squares deviation between model Drowning Incidences and GBD Incidences Baseline
  - Payoff 2: Least squares deviation between model Drowning Deaths and GBD Deaths Baseline
  - Both payoffs equally weighted
- **Number of Optimization Runs:** Not explicitly reported (software default stopping criteria: convergence tolerance or maximum iterations = 10,000)
- **Final Calibrated Parameters:** See Section 3.3 above

**No sensitivity analysis reported in this scoping study** - Infrastructure exists (IP1-IP6, ES1-ES6) but not systematically explored.

### 5. IMPLEMENTATION

#### 5.1 Software or programming language

**Location in manuscript:** Methods (Data and Implementation); Reference 14

#### Response:

- **Software:** Stella Architect (isee systems, inc.)
- **Version:** 4.0
- **Operating System:** Windows
- **License:** Commercial software (educational/research license)

#### Specialized Stocks Used: None

- Model uses only standard SD stocks (rectangular accumulations)
- No conveyors, queues, ovens, or other specialized delay structures employed

**Reference:** STELLA Architect. Version 4.0. isee systems, inc.; 2025. [https://www.iseesystems.com](https://www.iseesystems.com/)

#### 5.2 Random sampling Location in manuscript: N/A

**Response:** Not applicable - This is a deterministic model with no stochastic elements, random sampling, or Monte Carlo methods.

#### 5.3 Model execution

**Location in manuscript:** Methods (Data and Implementation); Stella model file metadata

#### Response:

- **Integration Method:** Euler (first-order explicit)
- **Time Step (DT):** 0.0625 years
- **Simulation Duration:** 50 years (1990-2040)
- **Number of Time Steps:** 800 steps (50 / 0.0625)

**Rationale for DT=0.0625:** Selected to ensure numerical stability given the fastest time constant in the model (Perception Adjustment Rate = 0.995/year, implying ~1-year adjustment time). Time step represents ~1/16 of fastest time constant, well within stability bounds for Euler integration.

#### 5.4 System Specification Location in manuscript: N/A

**Response:**

- **Model Run Time:** <1 second (instantaneous for single deterministic run)
- **Hardware:** Not critical for this model (runs on standard desktop/laptop computers)
- **No special computing requirements:** Model is computationally lightweight (5 stocks, ~30 auxiliary variables, 50-year simulation)

**Note:** Run time would increase if sensitivity analysis or optimization is performed (multiple model runs), but base case execution is trivial.

### 6. CODE ACCESS

#### 6.1 Computer Model Sharing Statement

**Location in manuscript:** Data Availability Statement; Acknowledgements

#### Response:

**Model Availability:**

- Full Stella Architect model file (.stmx format) available upon request
- Model cannot be executed without Stella Architect software (commercial license required)

#### Data Availability:

- GBD data publicly available: Global Burden of Disease Study 2021, Institute for Health Metrics and Evaluation (IHME), Seattle, WA, USA
- GBD data access: <https://vizhub.healthdata.org/gbd-results/>
- Specific Ohio drowning mortality data (1990-2021) used for calibration available in published GBD results portal

#### Reproducibility:

- To reproduce results, researchers need:

1. Stella Architect 4.0 or compatible version (commercial software, ~$500-1500 depending on license)
2. GBD 2021 drowning mortality data for Ohio (publicly available)
3. Model file from authors (available upon request)

#### Limitations to Sharing:

- No public repository (e.g., GitHub) currently hosts the model
- Model file proprietary format (.stmx) requires Stella software to open
- Authors commit to sharing upon reasonable request to facilitate replication and extension

**Future Plans:** Authors invite injury prevention collaborators to request model files for adaptation to other jurisdictions (see Acknowledgements section).

### CHECKLIST COMPLETION STATEMENT

All applicable STRESS-SD items have been addressed. Items marked N/A reflect the scope of this study as a scoping model for behavior reproduction and hypothesis testing, not a policy analysis or optimization study. The model serves as a foundation for future research that will include:

- Multi-state validation and calibration
- Disaggregation by demographic factors (race, ethnicity, SES)
- Policy scenario analysis using intervention parameters (IP1-IP6)
- Sensitivity analysis across parameter ranges
- Optimization for equity-focused intervention strategies

**Completed by:** Brian J. Biroscak, PhD, MS, MA; Robinson Salazar Rua, PhD

**Date:** December 30, 2025

**Manuscript:** US Pediatric Drowning Trends: A System Dynamics Scoping Model Based on Global Burden of Disease (GBD) Estimates
